# Identifying circulating protein targets for common factors underlying schizophrenia, depression, and bipolar disorder

**DOI:** 10.64898/2026.06.01.26354643

**Authors:** Jiaqi Duan, Chen-Yang Su, Satoshi Yoshiji, Wenmin Zhang, Tianyuan Lu

## Abstract

**Background:** Schizophrenia, bipolar disorder, and depression share substantial genetic liability. However, the molecular mechanisms underlying this shared architecture remain poorly characterized. In particular, the role of circulating proteins as potential mediators and therapeutic targets is not well understood.

**Methods:** Based on large-scale genome-wide association studies, we constructed a latent psychiatric common factor using genomic structural equation modeling. We then performed proteome-wide Mendelian randomization to estimate the associations between circulating proteins and this shared liability, based on four independent proteomic cohorts. Protein-psychiatric common factor associations were prioritized through comprehensive sensitivity analyses and colocalization. We additionally performed tissue- and single-cell expression enrichment analyses and a systematic druggability assessment.

**Results:** We identified 36 circulating proteins with evidence of association with the psychiatric common factor that withstood multiple sensitivity analyses. Several proteins showed distinct tissue-specific expression patterns, with enrichment in brain, immune, or liver tissues, highlighting convergent neuroimmune and systemic pathways. For instance, genetically predicted higher levels of MAPK3, FES, MRE11A, HS6ST3, OLFM1, BTN3A1, BTN3A2 and BTN3A3 were associated with increased psychiatric risk, whereas higher levels of CD40, ITIH3, and ITIH4 were associated with decreased risk. Druggability assessment identified CD40, MAPK3, FES, MRE11A and BTN3A1 as established or potential therapeutic targets.

**Conclusions:** By integrating genetic, proteomic, and transcriptomic data, this study identifies circulating proteins that associated with the shared genetic effects on three major psychiatric disorders. These findings provide biologically grounded candidates for therapeutic targeting and offer insights into shared disease mechanisms.

## Introduction

Psychiatric disorders incur a substantial global disease burden and are characterized by marked clinical and biological heterogeneity (1–3). Large-scale genome-wide association studies (GWAS) of major psychiatric disorders, including schizophrenia, depression, and bipolar disorder, have identified hundreds of associated loci, providing unprecedented opportunities to uncover biological mechanisms and identify potential biomarkers and therapeutic targets (4–6).

Proteins represent an intermediate molecular layer linking genetic variation to disease phenotypes and therefore offer a promising avenue for translating genetic findings into biological insight (7). Compared with genetic variants, proteins are functionally interpretable, directly involved in cellular pathways, and often measurable in accessible biological samples such as blood (8–10). Importantly, many proteins are pharmacologically tractable, making them attractive candidates for biomarker discovery and drug development (11). Circulating proteins are particularly informative because they can reflect systemic physiological processes, including immune activity, metabolic regulation, and tissue injury, and can be readily quantified and potentially modulated through therapeutic interventions (12,13).

One promising strategy to investigate the roles of circulating proteins in complex diseases is Mendelian randomization (MR) using genetic variants associated with protein abundance as instrumental variables (14–20). Recent large-scale proteogenomic studies have identified numerous robust protein quantitative trait loci (pQTLs), enabling systematic evaluation of protein–disease relationships (21–24). In particular, *cis*-pQTLs that are more likely to influence protein levels through local regulatory mechanisms provide strong instruments for MR analyses (14). When core MR assumptions are satisfied, this approach can provide estimates of the effects of circulating proteins on disease risk while mitigating confounding and reverse causation (25).

Using pQTL-facilitated MR, previous studies, including our own work, have begun to investigate the roles of circulating proteins in psychiatric disorders (15,26). In earlier analyses examining individual disorders, we identified several proteins with potential effects on psychiatric disease risk. For example, genetically increased levels of TIMP4 were associated with a reduced risk of bipolar disorder, whereas genetically increased levels of ESAM were associated with an increased risk of schizophrenia (19). These findings highlight the potential of circulating proteomics to uncover biologically meaningful biomarkers and therapeutic targets for psychiatric disorders.

However, these prior analyses considered each disorder in isolation, despite substantial evidence that many psychiatric conditions share a common genetic architecture (27,28). Structural equation modeling provides a natural framework to account for this shared liability by explicitly modeling the covariance among disorders (29). Since many psychiatric disorders share substantial genetic liability, overlapping biological pathways may contribute to their pathogenesis, raising the possibility that some circulating proteins influence shared biological mechanisms underlying multiple conditions (30,31). Identifying such proteins could have important implications, as they may represent broadly relevant biomarkers or therapeutic targets capable of impacting multiple disorders simultaneously.

Another key question concerns the biological origins of circulating proteins that show evidence of potential effects on psychiatric disease risk. Circulating proteins can arise from diverse tissues and cell types (32). Characterizing their primary tissues of expression may provide important clues regarding the biological pathways through which they act. In particular, enrichment of candidate proteins in brain tissues may suggest direct involvement in neural processes, whereas enrichment in immune tissues may reflect neuroimmune mechanisms that influence brain function (33). In addition, the liver produces a large proportion of circulating proteins and plays a central role in systemic metabolism and inflammation (34). Identifying liver-expressed proteins may help clarify systemic pathways linking peripheral physiology to psychiatric disease risk.

Therefore, in this study, we aimed to systematically identify circulating proteins that influence the shared genetic liability underlying major psychiatric disorders. We leveraged the largest available GWAS of schizophrenia, depression, and bipolar disorder, and applied genomic structural equation modeling (GenomicSEM) to construct a latent common factor capturing their shared genetic architecture (28,35–37). We then conducted proteome-wide Mendelian randomization (MR) analyses using protein instruments derived from four large pQTL studies, encompassing the Atherosclerosis Risk in Communities (ARIC) study, the deCODE genetics study, the Fenland study, and the UK Biobank Pharma Proteomics Project (UKB-PPP) (21–24).To ensure robustness, we performed multiple sensitivity analyses to assess potential violations of MR assumptions. We further characterized prioritized proteins by examining tissue-and cell-type–specific gene expression patterns using bulk and single-cell transcriptomic datasets. Finally, we evaluated the druggability of candidate proteins using multiple pharmacogenomic databases. Through this integrative framework, our study aims to identify circulating proteins that may serve as biomarkers or therapeutic targets influencing the shared genetic architecture of schizophrenia, depression, and bipolar disorder.

## Methods and Materials

### Genome-wide association studies

GWAS summary statistics for schizophrenia, depression and bipolar disorder were obtained from the Psychiatric Genomics Consortium (PGC). Analyses were restricted to individuals of European ancestry due to the limited sample sizes of non-European ancestry cohorts. The European ancestry GWAS included 53,386 cases and 77,258 controls from 76 cohorts for schizophrenia (35), 412,305 cases and 1,588,397 controls from 75 cohorts for depression (36), and 59,287 cases and 781,022 controls from 68 cohorts for bipolar disorder (37).

Across cohorts, cases were defined using standardized but heterogeneous ascertainment procedures, including structured or semi-structured clinical interviews, clinician-administered diagnoses, medical records, registry data, questionnaire-based assessments, and validated self-report measures, according to established diagnostic criteria such as DSM-IV/DSM-5 or ICD-10/ICD-11 where applicable (35–37).

Genome-wide association analyses were conducted in cohorts with individual-level genotype data following standardized procedures developed by the PGC, including rigorous technical and genomic quality control. Inverse-variance weighted fixed-effects meta-analyses were conducted based on cohort-level summary statistics.

### Common factor analysis

Summary statistics were harmonized across studies to ensure consistency in genome build, allele coding, and variant identifiers. Standard quality control procedures were applied, including filtering for variants with a minor allele frequency > 0.01 and removal of strand-ambiguous variants. Variants were aligned to the 1000 Genomes Phase 3 European ancestry reference panel (38), and alleles were oriented consistently across all disease outcomes.

Genetic covariance structure was estimated using linkage disequilibrium score regression (LDSC) (39). Univariate LDSC was first used to estimate heritability for each disease outcome, followed by bivariate LDSC to estimate pairwise genetic correlations among schizophrenia, depression, and bipolar disorder. These estimates were assembled into a genetic covariance matrix and its associated sampling covariance matrix, which served as inputs for subsequent structural equation modeling.

A common factor model was specified within the GenomicSEM R package (version 0.0.5) to capture shared genetic liability across the three psychiatric disorders (28,40). A single latent common factor was modeled as underlying schizophrenia, depression, and bipolar disorder, with factor loadings freely estimated and the latent common factor variance fixed to 1 for identification. Model parameters, including the magnitude of the association between each variant and the common factor, were estimated using a diagonally weighted least squares estimator, which accounts for uncertainty in the estimated genetic covariance matrix.

### Circulating protein quantitative trait locus studies

To comprehensively assess the potential effect of the plasma proteome on the common factor GWAS, we utilized genetic associations from four large-scale proteomic GWAS derived from individuals of European ancestry (21–24). These included the ARIC study, the deCODE genetics study, the Fenland study, and the UKB-PPP. In each cohort, circulating protein abundances were quantified in plasma samples, followed by genome-wide association analyses to identify genetic variants associated with protein levels.

The ARIC, deCODE, and Fenland study quantified proteins using the SomaScan platform based on Slow Off-rate Modified Aptamers (SOMAmer), whereas UKB-PPP employed the antibody-based Olink Proximity Extension Assay (PEA) technology (21–24). Although differences existed across studies in measurement platforms and normalization procedures, protein levels were standardized within each cohort such that genetic effect estimates could be interpreted as allele effects corresponding to one standard deviation (SD) change in protein abundance, thereby ensuring comparability of effect sizes across cohorts.

### Genetic instrument selection and two-sample Mendelian randomization analysis

Using GWAS summary statistics of the psychiatric common factor, we conducted two-sample MR analyses to evaluate the associations between genetically predicted circulating protein levels and shared psychiatric genetic liability.

We defined genetic instruments for circulating protein levels based on *cis*-pQTLs defined as variants located within 500 kb of the transcription start site (TSS) of the encoding gene. Independence was defined using a linkage disequilibrium (LD) threshold of r^2^ < 0.001, window size of 1 Mb, and *P* < 5×10^−8^. To mitigate the risk of horizontal pleiotropy, we prioritized “strict V2G *cis*-pQTLs” supported by strong Variant-to-Gene (V2G) evidence as primary instruments following our previous work (41). When a *cis*-pQTL was unavailable in the outcome GWAS, proxy variants in high LD (r^2^ > 0.8) were identified using the 1000 Genomes European ancestry reference panel (38). Palindromic variants with high minor allele frequency (> 0.42) were excluded to avoid strand misalignment.

We performed two-sample MR to estimate the association of each protein with the common factor. For proteins with a single valid instrumental variable, the Wald ratio method was used. For proteins with two or more independent instruments, we employed the inverse-variance weighted (IVW) approach as the primary analysis. For proteins with at least three instruments, weighted median, penalized weighted median, and weighted mode methods were additionally performed as sensitivity analyses (25,42). Statistical significance was determined using a genome-wide suggestive threshold (*P* < 1×10^−5^; corresponding to a Bonferroni-corrected threshold for approximately 5,000 tests) to account for multiple testing.

All MR analyses were conducted using the TwoSampleMR R package (version 0.5.6).

### Sensitivity analyses and colocalization assessment

To assess the robustness of our findings and address potential violations of MR assumptions, we conducted a series of sensitivity analyses. Associations were considered robust only if they satisfied all predefined criteria. Specifically, for proteins instrumented by multiple variants, heterogeneity was evaluated using Cochran’s Q test and the *I^2^* statistic; associations showing significant heterogeneity (Cochran’s Q *P* < 0.05 and I^2^ ≥ 0.5) were excluded. Directional pleiotropy was assessed using the MR-Egger intercept (43), and associations were removed if the intercept deviated from zero (*P* < 0.05).

We further required consistency in the direction of effect across all cohort and MR methods, retaining only associations with consistent effect direction across all contributing cohorts for which effect estimates from Wald ratio or IVW, weighted median, weighted mode, and MR-Egger analyses were directionally concordant. In addition, Steiger filtering was applied to verify the direction of effect, excluding instances in which genetic instruments explained more variance in the outcome than in the exposure (44).

To distinguish putatively effects from confounding due to linkage disequilibrium, we performed colocalization analysis using SharePro with default prior settings (45,46). A posterior probability of colocalization ≥ 0.8 was considered evidence of a shared genetic association between the protein and the outcome.

### Tissue-specific expression profiling

To systematically evaluate the tissue expression patterns of prioritized protein-coding genes, we analyzed transcriptomic data from the Genotype-Tissue Expression project version 8 (GTEx v8), which provides gene expression profiles across 54 non-diseased human tissues (47). This dataset includes RNA sequencing data from approximately 1,000 individuals. We retained genes that were detected (at least 5 reads) in at least 20% of samples within each tissue. Within each tissue, gene expression levels were normalized using the trimmed mean of M-values (TMM) method implemented in the edgeR package (48). For each gene, we then calculated the median TMM value across individuals within each tissue to represent its overall expression level in that tissue (49). To characterize tissue-specific expression patterns, we applied a two-step robust standardization procedure. First, within each tissue, gene-level median expression values were standardized using the median and median absolute deviation (MAD) across all genes. Second, for each gene, the within-tissue standardized values were further normalized across tissues using the median and MAD calculated across tissues, yielding tissue-specific enrichment z-scores.

We focused on three biologically relevant groups of tissues: (1) brain tissues included amygdala, anterior cingulate cortex (BA24), caudate (basal ganglia), cerebellar hemisphere, cerebellum, cortex, frontal cortex (BA9), hippocampus, hypothalamus, nucleus accumbens (basal ganglia), putamen (basal ganglia), spinal cord (cervical c-1), and substantia nigra; (2) immune-related tissues included spleen, whole blood, and Epstein Barr virus (EBV)-transformed lymphocytes; and (3) liver. Brain tissues were examined because enrichment in neuronal or glial populations may suggest direct involvement in neural processes relevant to psychiatric disease pathophysiology (50,51). Immune tissues were analyzed given growing evidence that neuroimmune interactions contribute to the development and progression of psychiatric disorders (52). In addition, the liver was evaluated because it produces a substantial proportion of circulating proteins and plays a central role in systemic metabolic and inflammatory processes that may influence brain function (53).

A threshold of z > 10 was used to define significant expression enrichment in a given tissue (18). Genes exhibiting significant enrichment in brain tissues, immune tissues, or liver, while not showing higher enrichment z-scores in other tissues, were retained as prioritized protein-coding genes for downstream analyses.

### Single-cell gene expression profiling across brain, immune, and liver tissues

We further investigated whether the genes encoding prioritized proteins exhibited expression enrichment in specific brain, immune, and liver cell types to better understand the potential biological origins and mechanisms underlying their circulating levels.

For immune tissues, we utilized a cross-tissue single-cell RNA sequencing dataset comprising 329,762 high-quality immune cells derived from 12 donors (54).

For brain tissues, we used the human single-cell/single-nucleus RNA sequencing dataset from the Allen Brain Cell Atlas (ABC Atlas) (55). This dataset is based on single-nucleus transcriptomic profiles from multiple brain regions and comprises over 3,000,000 cells and nuclei.

We also utilized a publicly available single-cell RNA sequencing dataset with GEO accession GSE124395, which generated transcriptomic profiles from approximately 10,000 high-quality cells derived from normal human liver tissue (56).

We evaluated the normalized expression levels of each target protein-coding gene at the single-cell level and compared the distribution of gene expression across different annotated cell types.

### Druggability assessment

We performed a systematic druggability assessment for all prioritized proteins by querying multiple curated pharmacogenomic databases. Target proteins were stratified into druggability tiers based on the framework described by Finan et al., which integrates evidence across multiple databases (57). Specifically, Tier 1 comprised targets of approved drugs and drugs in clinical development; Tier 2 included proteins closely related to known drug targets or those with associated drug-like compounds; and Tier 3 encompassed extracellular proteins and members of key drug target families.

Furthermore, using standardized gene symbols, the coding genes for the candidate proteins were systematically searched in DrugBank, ChEMBL, and DGIdb (Drug-Gene Interaction Database). DrugBank was used to obtain manually curated target annotations and information on approved and investigational drugs (58). ChEMBL was used to extract bioactivity-supported target-compound relationships and target classifications (e.g., enzyme, receptor, ion channel) (59). DGIdb was used to aggregate reported drug-gene interaction evidence from multiple curated resources and literature-mined sources (60).

## Results

### Common factor analysis

The study design is presented in Figure 1. Using LDSC, we confirmed significant positive genetic correlations between each pair of the three psychiatric disorders (Supplementary Table 1), with the strongest correlation observed between bipolar disorder and schizophrenia (r_g_ = 0.643, SE = 0.030), followed by bipolar disorder-depression (r_g_ = 0.520, SE = 0.026) and schizophrenia-depression (r_g_ = 0.357, SE = 0.023).

**Figure 1.**
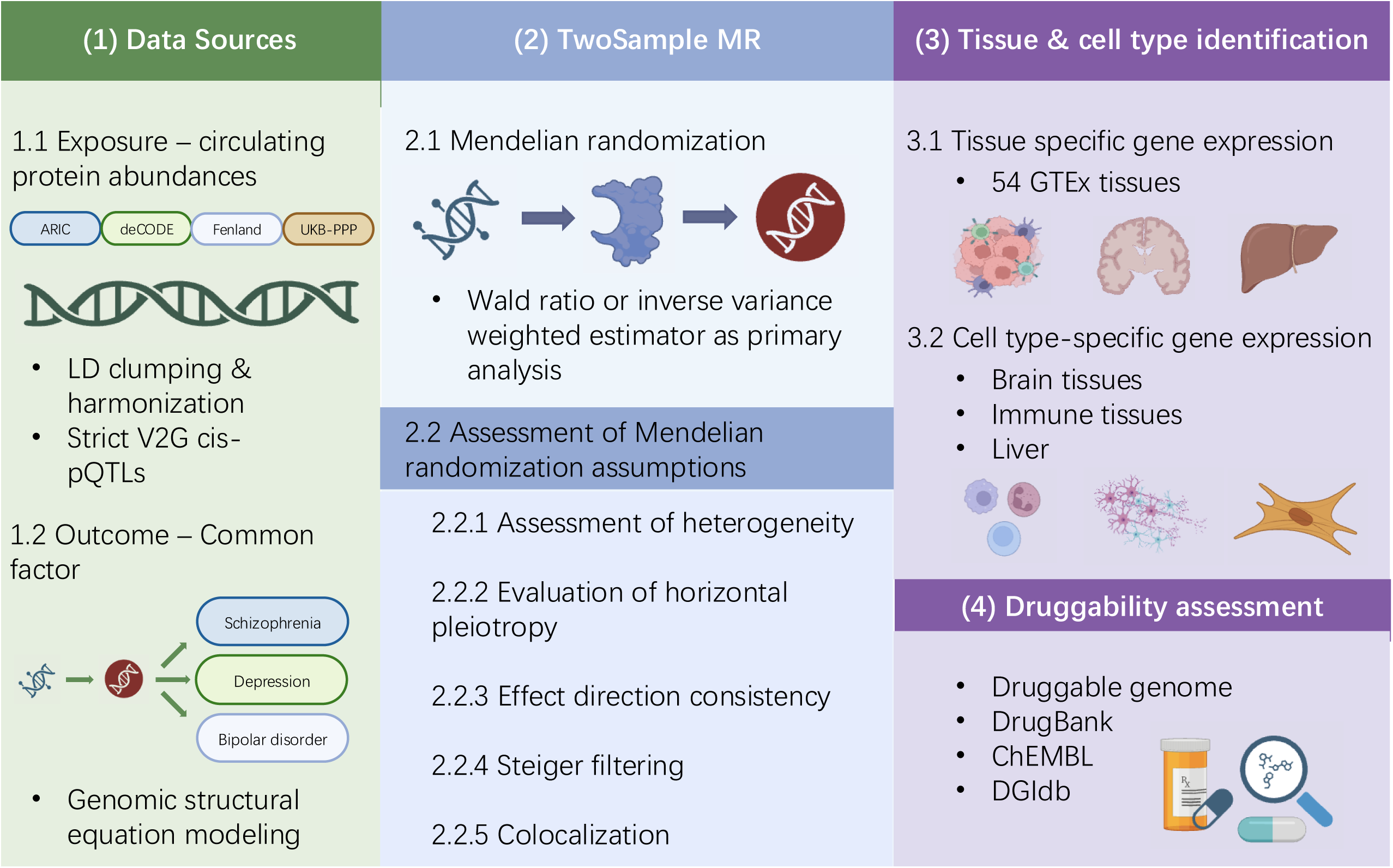
Overview of study. Common factor analysis was performed based on GWAS summary statistics for schizophrenia, bipolar disorder, and depression using Genomic SEM. Two-sample MR was conducted with circulating protein abundances from four proteomic GWAS (ARIC, deCODE, Fenland, and UKB-PPP) as exposures and the common factor as outcome. Multiple sensitivity analyses and colocalization analyses performed to ensure robustness and minimize confounding. Tissue- and cell type-specific expression analyses were used to identify relevant biological contexts. Druggability assessment was conducted to prioritize potential therapeutic targets.

Using GemonicSEM, we constructed a latent common factor capturing shared genetic liability across schizophrenia, depression, and bipolar disorder, and estimated genome-wide associations with this factor. As expected, the common factor exhibited a polygenic architecture (Figure 2), with an LDSC-estimated heritability of 0.137 (SE = 0.004). The LDSC intercept was 0.999 (SE = 0.013), indicating a low risk of residual confounding (39).

**Figure 2.**
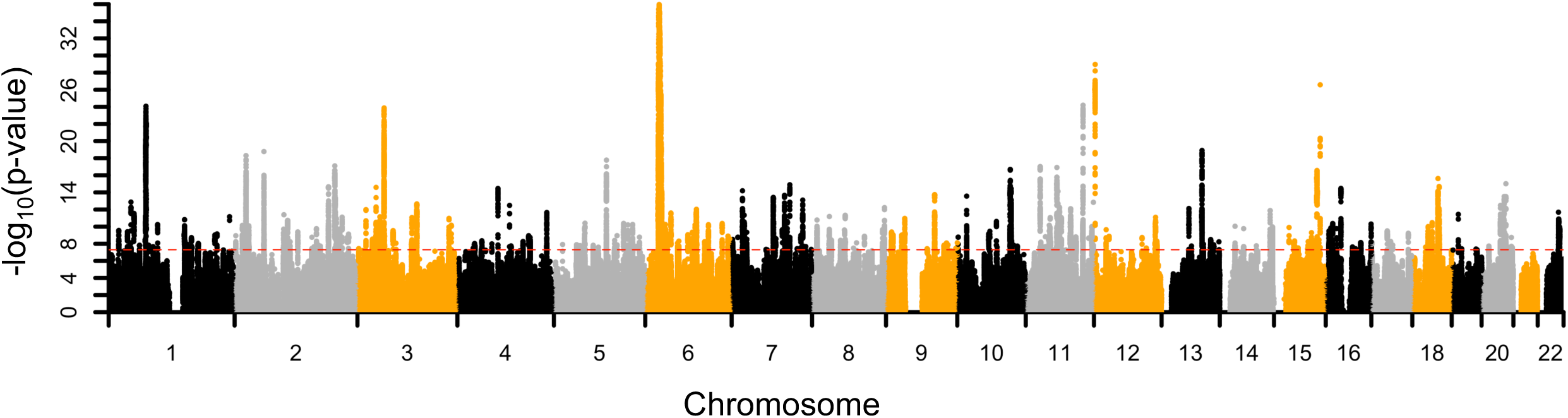
Manhattan plot illustrating genome-wide associations with the common factor. The x-axis represents the genomic positions of variants, and the y-axis shows the corresponding −log_10_(p-values). The red dashed line indicates the genome-wide significance threshold (P = 5×10^−8^).

### Proteome-wide MR

Using four large-scale pQTL studies conducted in individuals of European ancestry (ARIC, deCODE, Fenland, and UKB-PPP), we systematically evaluated the potential effects of circulating proteins on the common factor. The numbers of proteins tested were 1,507 based on ARIC, 1,602 based on deCODE, 1,422 based on Fenland, and 1,579 based on UKB-PPP, respectively.

A total of 46 circulating protein-common factor associations reached the genome-wide suggestive threshold *(P* < 1 × 10^−5^; Supplementary Table 2). Of these associations, 36 withstood sensitivity analyses and colocalization analyses, and demonstrated consistent effect direction across contributing cohorts (Supplementary Figure 1 and Supplementary Table 3). Among these proteins, ERBB4 was instrumented by three or more independent *cis*-variants. For this protein, effect estimates derived from the weighted median, penalized weighted median, weighted mode, and MR-Egger methods were highly consistent with those obtained from the IVW approach. All instruments used for these tests passed Steiger filtering.

The direction and magnitudes of these associations are illustrated in Figure 3 and Supplementary Figure 2. Specifically, genetically predicted increases in circulating levels of MAPK3, FES, MRE11A, HS6ST3, OLFM1, BTN3A1, BTN3A2, BTN3A3, and PC were associated with higher values of the common factor. The estimated effects of a one standard deviation increase in circulating protein levels on the common factor were 0.10 (95% CI: 0.06 to 0.13) for MAPK3, 0.28 (95% CI: 0.22 to 0.34) for FES, 0.19 (95% CI: 0.11 to 0.28) for MRE11A, 0.13 (95% CI: 0.08 to 0.19) for HS6ST3, 0.12 (95% CI: 0.07 to 0.17) for OLFM1, 0.15 (95% CI: 0.11 to 0.19) for BTN3A1, 0.08 (95% CI: 0.07 to 0.10) for BTN3A2, 0.04 (95% CI: 0.02 to 0.05) for BTN3A3, and 0.23 (95% CI: 0.15 to 0.30) for PC.

**Figure 3.**
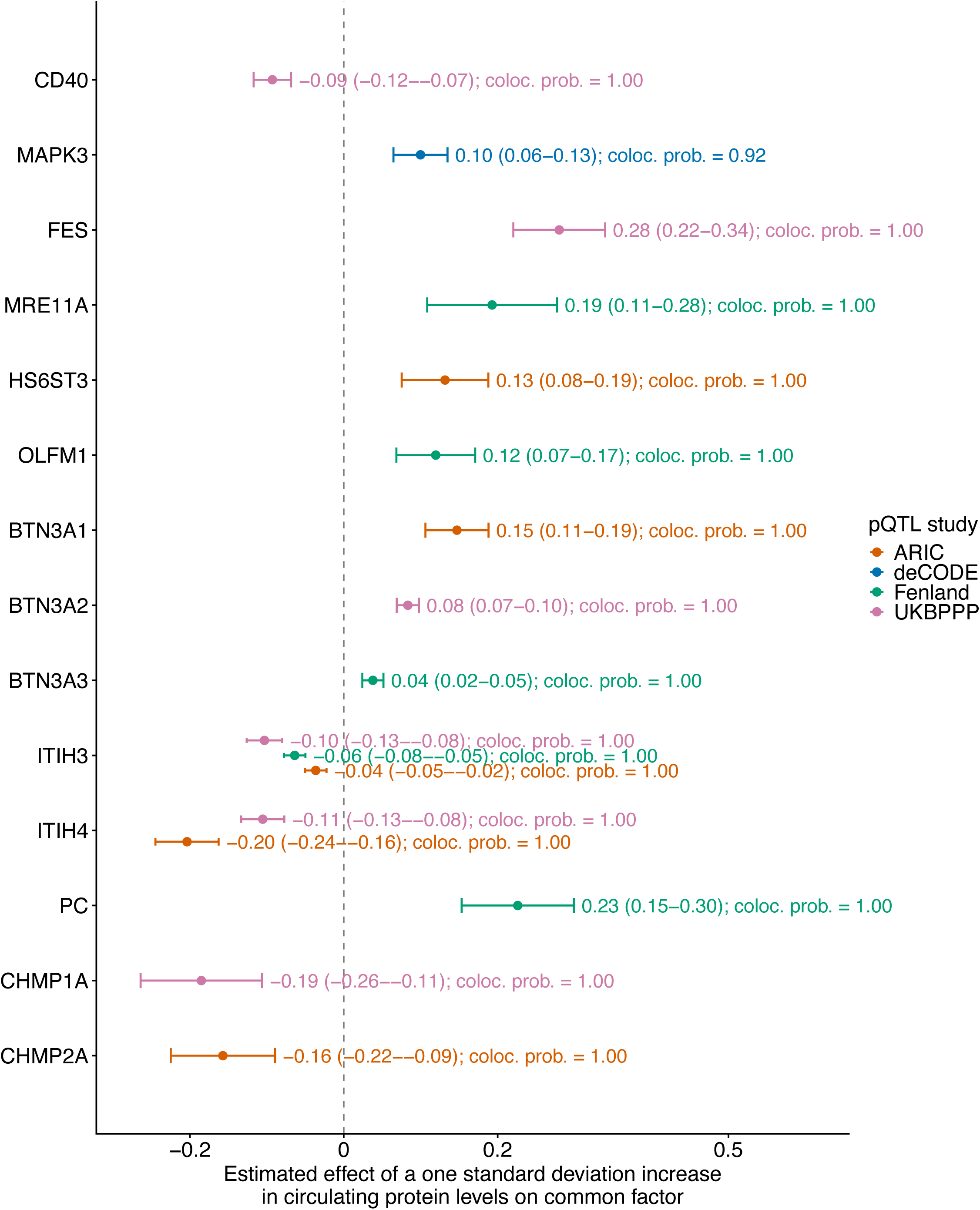
Illustration of estimated associations between circulating protein levels and common factor. Associations that were significant in MR analyses, withstood sensitivity analyses and colocalization analyses, and demonstrated enrichment in brain, immune, or liver tissues are illustrated. The remaining associations are illustrated in Supplementary Figure 2. Each dot denotes to the estimated effect of a protein based on a specific pQTL study, with error bars indicating 95% confidence intervals. Colocalization probabilities (coloc. prob.) are indicated. The vertical dashed line denotes the null.

In contrast, genetically predicted increases in circulating levels of CD40, ITIH3, ITIH4, CHMP1A, and CHMP2A were associated with lower values of the common factor. A one standard deviation increase in genetically predicted circulating protein levels was associated with a decrease of 0.09 (95% CI: −0.12 to −0.07) for CD40, up to 0.10 (95% CI: −0.13 to −0.08) for ITIH3, up to 0.20 (95% CI: −0.24 to −0.16) for ITIH4, 0.19 (95% CI: −0.26 to −0.11) for CHMP1A, and 0.16 (95% CI: −0.22 to −0.09) for CHMP2A in the common factor (Figure 3, Supplementary Figure 2 and Supplementary Table 3).

For TIMP4 and ESAM, we obtained effect estimates that were directionally consistent with those reported in our previous study focusing on individual disorders (15). Specifically, a one standard deviation increase in genetically predicted TIMP4 was associated with a decrease of up to 0.05 (95% CI: −0.07 to −0.03) in the common factor, while ESAM was associated with an increase of up to 0.12 (95% CI: 0.07 to 0.16).

### Bulk tissue and single cell enrichment of gene expression

For each target protein-coding gene, we evaluated tissue-specific expression enrichment across 54 tissue sites using data from GTEx to identify candidate tissues (Supplementary Table 4). As a result, *HS6ST3* and *OLFM1* exhibited enrichment in brain tissues. Additionally, *MAPK3, FES, MRE11A, BTN3A1*, *BTN3A2*, *BTN3A3*, *CHMP2A*, *CHMP1A*, and *CD40,* were enriched in immune-related tissues, while *ITIH3*, *ITIH4*, and *PC* were enriched in liver. Tissue-specific gene expression patterns are shown in Figure 4 and Supplementary Table 4.

**Figure 4.**
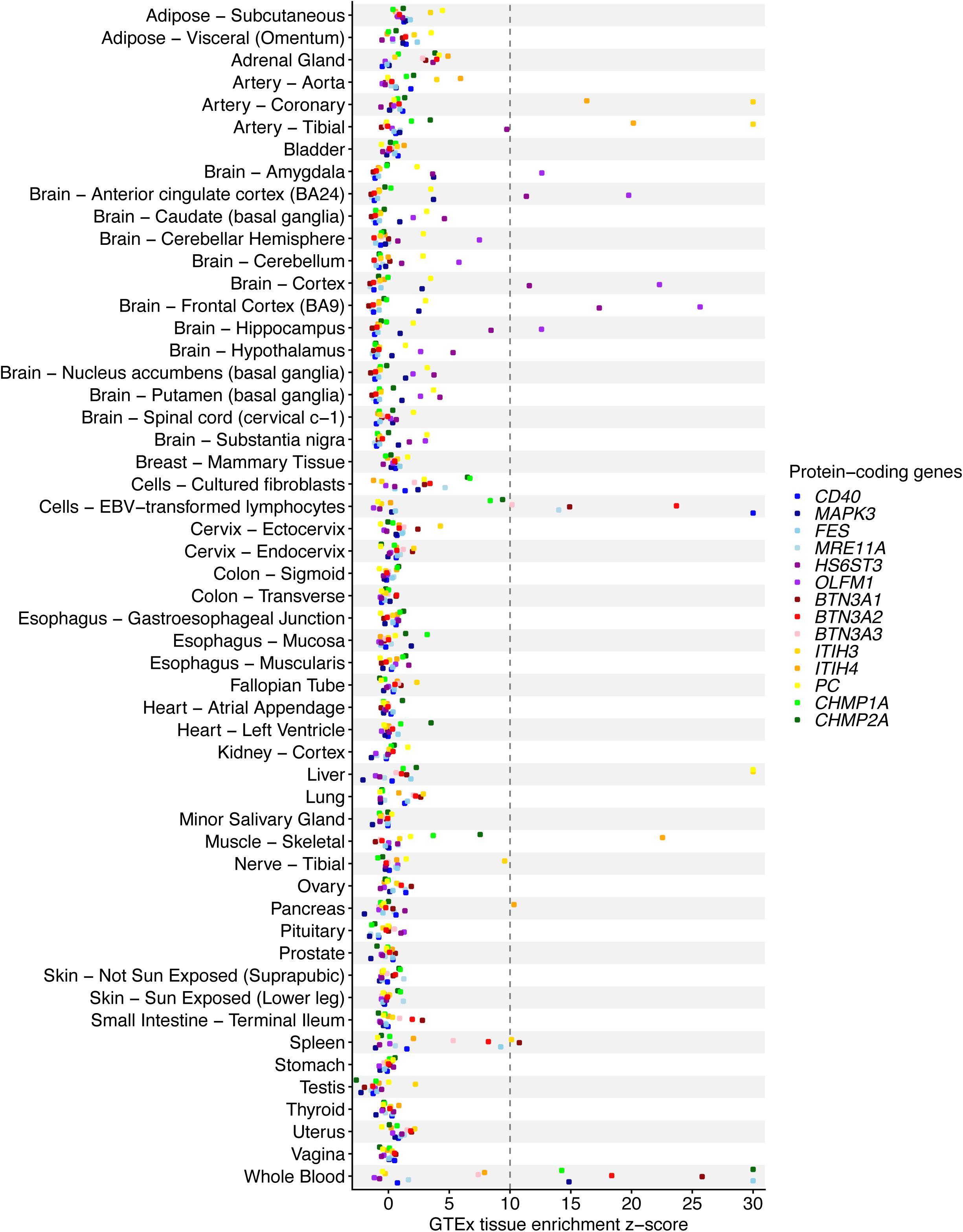
Tissue-specific expression patterns of prioritized genes. Gene expression profiles were evaluated using GTEx v8 data across 54 tissues. Each dot denotes the z-score of a protein in a given tissue. The dashed vertical line marks the predefined enrichment threshold (z-score > 10). To improve visualization, z-scores were capped at 30.

We further examined single-cell transcriptomic profiles to characterize the cell type-specific expression patterns of prioritized genes enriched in brain, immune, and liver tissues.

In brain tissues, *HS6ST3* exhibited widespread expression across multiple excitatory neuronal subtypes and several inhibitory neuronal populations. In addition, elevated expression was observed in oligodendrocyte precursor cells, Bergmann glia, and choroid plexus cells. In contrast, OLFM1 exhibited broad and relatively high expression across multiple neuronal populations, particularly in hippocampal, cortical intratelencephalic, amygdala excitatory, and thalamic excitatory neurons, while showing limited expression in non-neuronal cell types. Brain tissue-specific single-cell gene expression patterns are shown in Supplementary Figure 3 and Supplementary Table 5.

In immune cell populations, several genes showed distinct lineage-specific expression patterns (Figure 5a, Supplementary Figure 4 and Supplementary Table 5). Members of the *BTN3A* family (*BTN3A1*, *BTN3A2*, *BTN3A3*) were enriched in T cells and natural killer (NK) cells, particularly within memory T-cell subsets, while showing lower expression in B cells and most myeloid populations. *CD40* displayed strong enrichment in B cells and was also expressed in selected antigen-presenting cells, whereas expression in T and NK cells was comparatively limited. In contrast, *FES* was predominantly enriched in the myeloid lineage, particularly in monocytes, macrophages, and dendritic cells. Several genes, including *MAPK3*, *MRE11A*, *CHMP2A*, and *CHMP1A*, showed relatively broad expression across immune cell types without clear lineage-specific enrichment.

**Figure 5.**
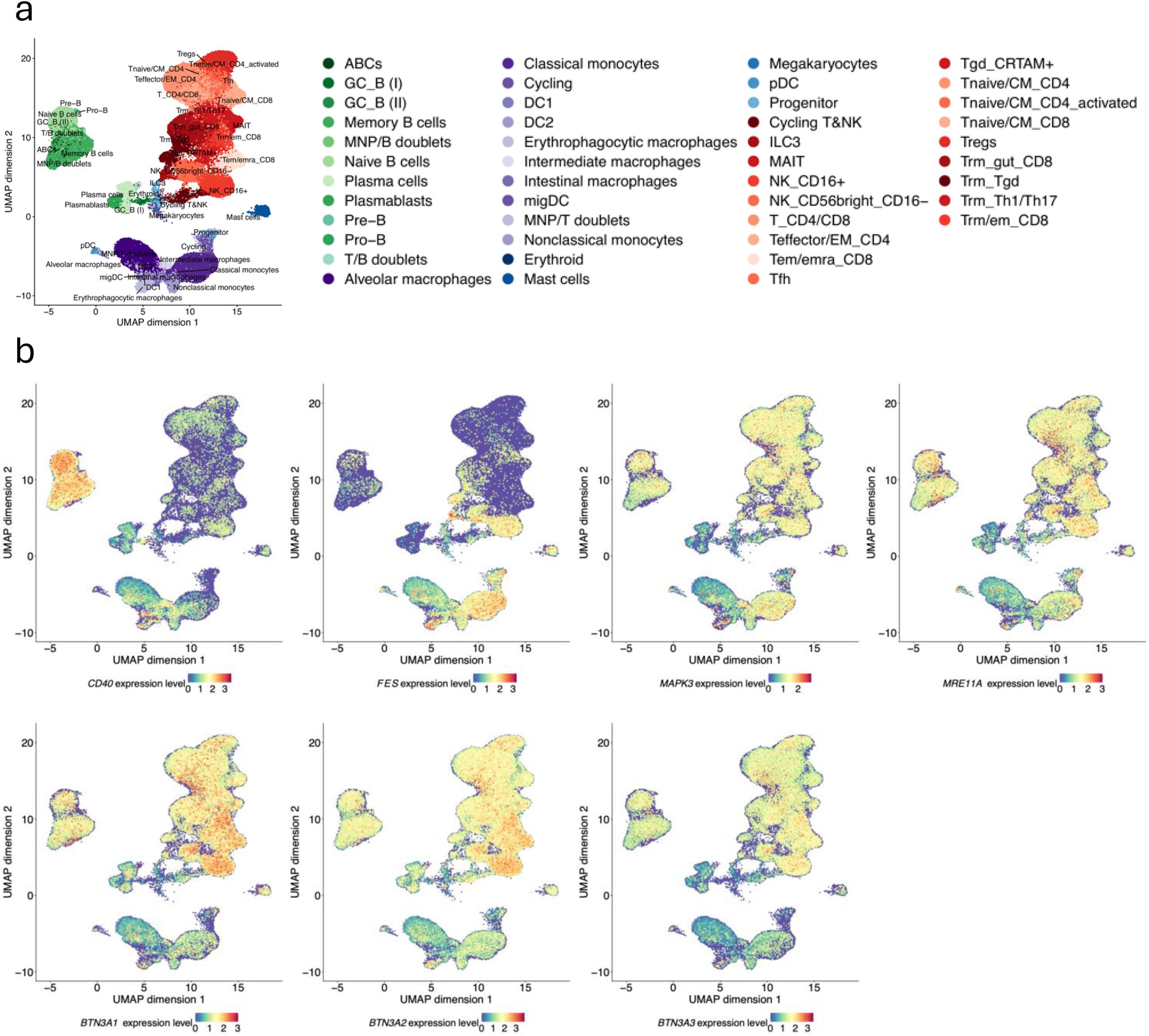
Single-cell gene expression patterns of selected genes across immune cell populations. Expression patterns were visualized using uniform manifold approximation and projection (UMAP) based on transcriptomic data from 329,762 cells. (a) Cell type annotations are illustrated. Each dot represents a single cell colored according to manually curated cell types. (b) Normalized gene expression levels of *CD40*, *FES*, *MAPK3*, *MRE11A*, *BTN3A1*, *BTN3A2*, and *BTN3A3* are shown.

In liver, members of the *ITIH* family (*ITIH3*, and *ITIH4*) and *PC* demonstrated pronounced hepatocyte-specific expression (Supplementary Figure 5 and Supplementary Table 5).

### Druggability assessment

We systematically integrated information from the druggable genome, DrugBank, ChEMBL, and DGIdb to evaluate the druggability of the 14 prioritized proteins. Among them, CD40, FES and BTN3A1 had documented drug–target interactions in at least one database, with several associated compounds having entered clinical development or received regulatory approval (Table 1). No drug-related records were identified for the remaining prioritized proteins.

**Table 1.**
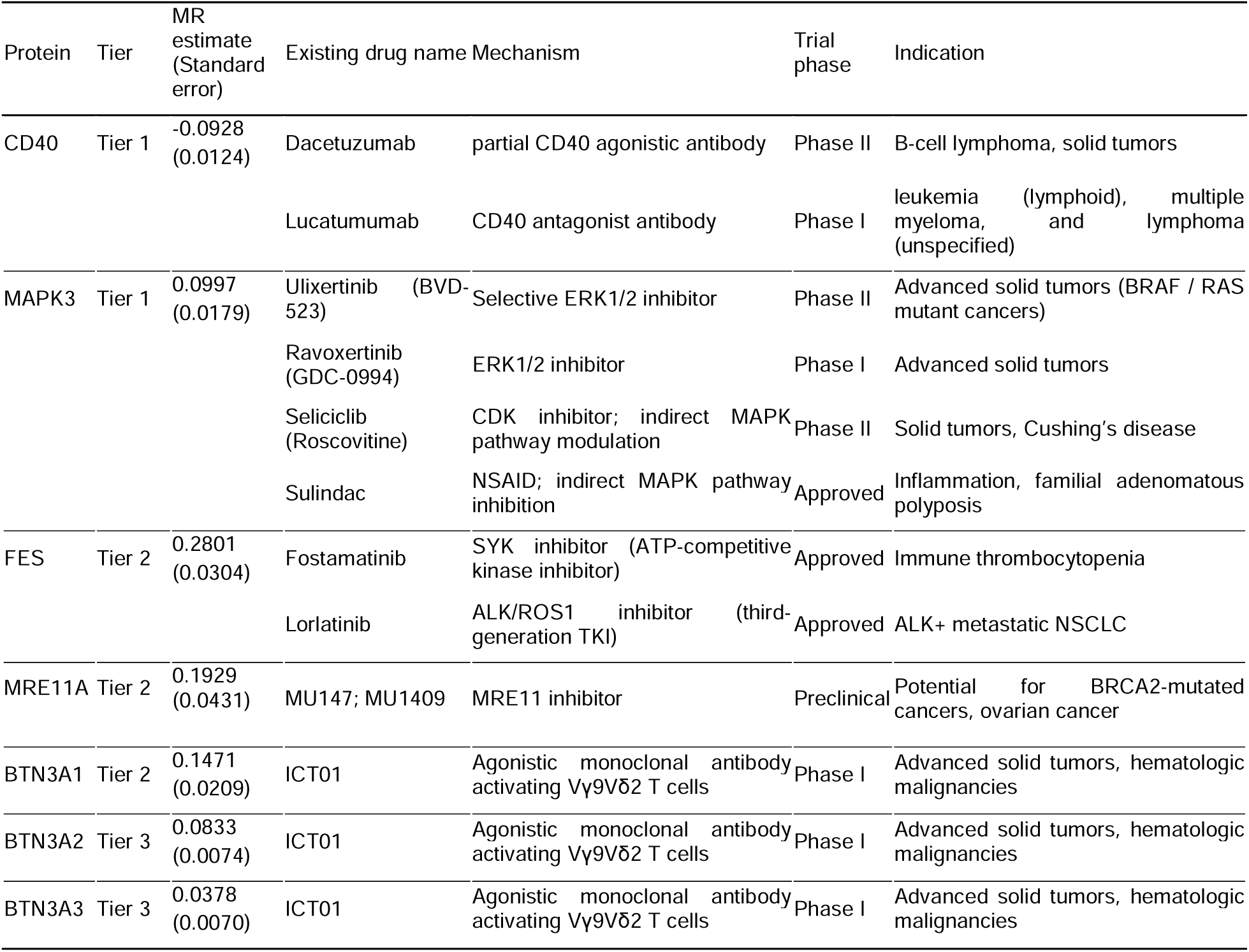
Druggability assessment of prioritized proteins.

CD40 was classified as a Tier 1 drug target and showed a negative effect estimate in the MR analysis. Two monoclonal antibodies targeting CD40 were identified: dacetuzumab, a partial agonistic antibody currently evaluated in phase II trials for B-cell lymphoma and solid tumors, and lucatumumab, an antagonistic antibody tested in phase I trials for lymphoid leukemia, multiple myeloma, and lymphoma.

MAPK3 (ERK1) was also categorized as a Tier 1 drug target and showed a positive effect in the MR analysis. Multiple compounds targeting the ERK/MAPK signaling pathway were identified, including the selective ERK1/2 inhibitor ulixertinib (BVD-523), currently in phase II trials for advanced solid tumors with BRAF or RAS mutations, and ravoxertinib (GDC-0994), evaluated in phase I trials for advanced solid tumors. Additional compounds with indirect modulation of the MAPK pathway include seliciclib (roscovitine), a cyclin-dependent kinase inhibitor evaluated in phase II trials for solid tumors and Cushing’s disease, and sulindac, a nonsteroidal anti-inflammatory drug approved for inflammatory conditions and familial adenomatous polyposis.

FES, MRE11A, and BTN3A1 were classified as Tier 2 drug targets and all exhibited a positive effect in the MR analysis. Two approved kinase inhibitors with reported interactions with FES were identified: fostamatinib, an ATP-competitive SYK inhibitor approved for immune thrombocytopenia, and lorlatinib, a third-generation ALK/ROS1 tyrosine kinase inhibitor approved for ALK-positive metastatic non-small cell lung cancer. Two preclinical small-molecule MRE11 nuclease inhibitors were identified: MU147 and MU1409, which have been investigated as potential therapeutic agents for DNA damage response–targeted cancer therapy, with studies reported in BRCA2-mutated cancers and ovarian cancer. The agonistic monoclonal antibody ICT01, which activates Vγ9Vδ2 T cells through the BTN3A family, has been evaluated in phase I clinical trials for advanced solid tumors and hematologic malignancies. Additional members of the BTN3A family, BTN3A2 and BTN3A3, were classified as Tier 3 targets and were also linked to ICT01 in the queried databases.

## Discussion

Psychiatric disorders are among the leading causes of disability worldwide and contribute substantially to premature mortality, impaired quality of life, and socioeconomic burden (61,62). Despite extensive genetic and epidemiological research, the biological mechanisms underlying their shared liability remain incompletely understood, limiting the development of effective biomarkers and targeted interventions. In this study, we integrated Genomic SEM, circulating proteomic pQTL data, Mendelian randomization (MR), single-cell transcriptomics, and druggability assessment to identify circulating proteins associated with the shared genetic architecture underlying schizophrenia, depression, and bipolar disorder, and to characterize their tissue and cellular expression patterns.

Our analyses identified multiple circulating proteins associated with the common genetic factor underlying schizophrenia, depression, and bipolar disorder. Integration of MR findings with single-cell transcriptomic enrichment and druggability assessment prioritized several biologically relevant candidates, including CD40, MAPK3, FES, MRE11A, HS6ST3, OLFM1, members of the BTN3A family, and members of the ITIH family. Collectively, these findings support the involvement of immune dysregulation, neuronal signaling, and extracellular matrix–related pathways in the shared biology of major psychiatric disorders (63–65).

Several prioritized proteins converged on immune-related mechanisms. CD40, MAPK3, FES, and members of the BTN3A family showed enrichment across immune cell populations, particularly within lymphoid and myeloid compartments. These findings are consistent with growing evidence implicating chronic immune activation, inflammatory signaling, and altered lymphocyte function in psychiatric disorders. CD40 is a key regulator of immune co-stimulation and inflammatory signaling through NF-κB, MAPK, and PI3K pathways, while MAPK3 is a central component of intracellular signaling networks involved in stress responses, inflammation, and synaptic regulation (66,67). FES has established roles in myeloid-cell signaling and inflammatory regulation, and BTN3A family members participate in phosphoantigen sensing and γδ T-cell activation (68,69). Together, these observations support the hypothesis that dysregulated peripheral immune signaling contributes to the shared genetic liability underlying psychiatric disorders.

In addition to immune-related pathways, several prioritized proteins highlighted central nervous system–related biological processes. HS6ST3 and OLFM1 showed enriched expression in neuronal populations and brain-related cell types, suggesting potential involvement in neuronal connectivity and synaptic regulation. HS6ST3 is involved in heparan sulfate sulfation and modulation of developmental and synaptic signaling pathways, whereas OLFM1 has been implicated in synaptic organization and neural connectivity (70,71). Although direct mechanistic evidence linking these proteins to psychiatric disorders remains limited, their brain-enriched expression profiles and consistent MR associations support their relevance to neurobiological processes underlying disease risk.

Our analyses also identified proteins linked to extracellular matrix and systemic regulatory pathways. Members of the ITIH family, particularly ITIH3 and ITIH4, demonstrated pronounced hepatocyte-specific expression and were associated with lower values of the shared genetic factor. The ITIH family is involved in extracellular matrix stabilization, inflammatory regulation, and maintenance of tissue microenvironments (72). Although previous observational studies have reported altered ITIH levels in psychiatric disorders, our findings suggest that these proteins may reflect broader systemic or compensatory biological processes related to psychiatric disease liability (73,74).

Beyond biological interpretation, several prioritized proteins have existing or emerging therapeutic relevance. CD40-, MAPK-, FES-, and BTN3A-related pathways are already being explored in immunology and oncology drug development, suggesting potential opportunities for future translational investigation in psychiatric disorders. At the same time, proteins enriched in brain tissues, such as HS6ST3 and OLFM1, may provide insight into disease mechanisms and contribute to future biomarker development or patient stratification strategies.

This study has several limitations. First, our analyses focused on schizophrenia, depression, and bipolar disorder because these disorders currently have the largest available GWAS sample sizes and the most robust genetic data. Future studies incorporating additional psychiatric and neurodevelopmental disorders may provide a more comprehensive understanding of the shared biological architecture across multiple disorders. Second, current circulating proteomic platforms provide incomplete coverage of the human proteome, and therefore potentially relevant circulating proteins may not have been captured in the present analyses. Third, circulating protein levels are not specific to brain tissues and may reflect systemic physiological or inflammatory processes rather than central nervous system-specific mechanisms alone. Fourth, the genetic and proteomic datasets were predominantly derived from individuals of European ancestry, which may limit the generalizability of these findings to other populations because of differences in allele frequencies, linkage disequilibrium patterns, and pQTL architecture (75,76). Fifth, although MR provides support for potential causal relationships, our analyses remain primarily statistical and do not include direct functional validation. Experimental studies will therefore be necessary to clarify the biological mechanisms through which prioritized proteins influence psychiatric disease risk. Sixth, additional lines of evidence are needed to corroborate these findings in future studies. In particular, large-scale longitudinal cohorts, deeply phenotyped psychiatric cohorts, and multi-omics datasets integrating proteomic, transcriptomic, and clinical information will be important for validating these associations and assessing their translational relevance. Finally, although we focused our interpretation on proteins showing enrichment in brain, immune, and liver tissues, other prioritized proteins may also be biologically relevant. Our prioritization strategy applied stringent significance and integrative evidence thresholds, and therefore additional proteins identified in our analyses warrant further investigation in future studies.

In conclusion, integrative proteogenomic analyses identified multiple circulating proteins associated with the shared genetic liability underlying schizophrenia, depression, and bipolar disorder. These findings support the involvement of immune, neuronal, and extracellular matrix–related pathways in the shared biology of psychiatric disorders and highlight several candidate proteins and signaling pathways for future mechanistic and translational investigation.

## Data Availability

All data produced in the present work are contained in the manuscript.

## Acknowledgements

Research reported in this publication was supported by the National Institutes of Health under Award Number R35GM162188. The content is solely the responsibility of the authors and does not necessarily represent the official views of the National Institutes of Health. T.L. has been supported by start-up funding from the Office of the Vice Chancellor for Research and Graduate Education, School of Medicine and Public Health, and Department of Population Health Sciences at the University of Wisconsin-Madison. The funders have no role in the conceptualization, design, data collection, analysis, decision to publish, or preparation of the manuscript. The authors gratefully acknowledge the Social Science Computing Cooperative at the University of Wisconsin-Madison for providing computing resources and statistical support.

## Consent

This study did not involve human participants, and therefore, informed consent was not required.

