## Supplementary Figures for "Identifying circulating protein targets for common factors underlying schizophrenia, depression, and bipolar disorder"

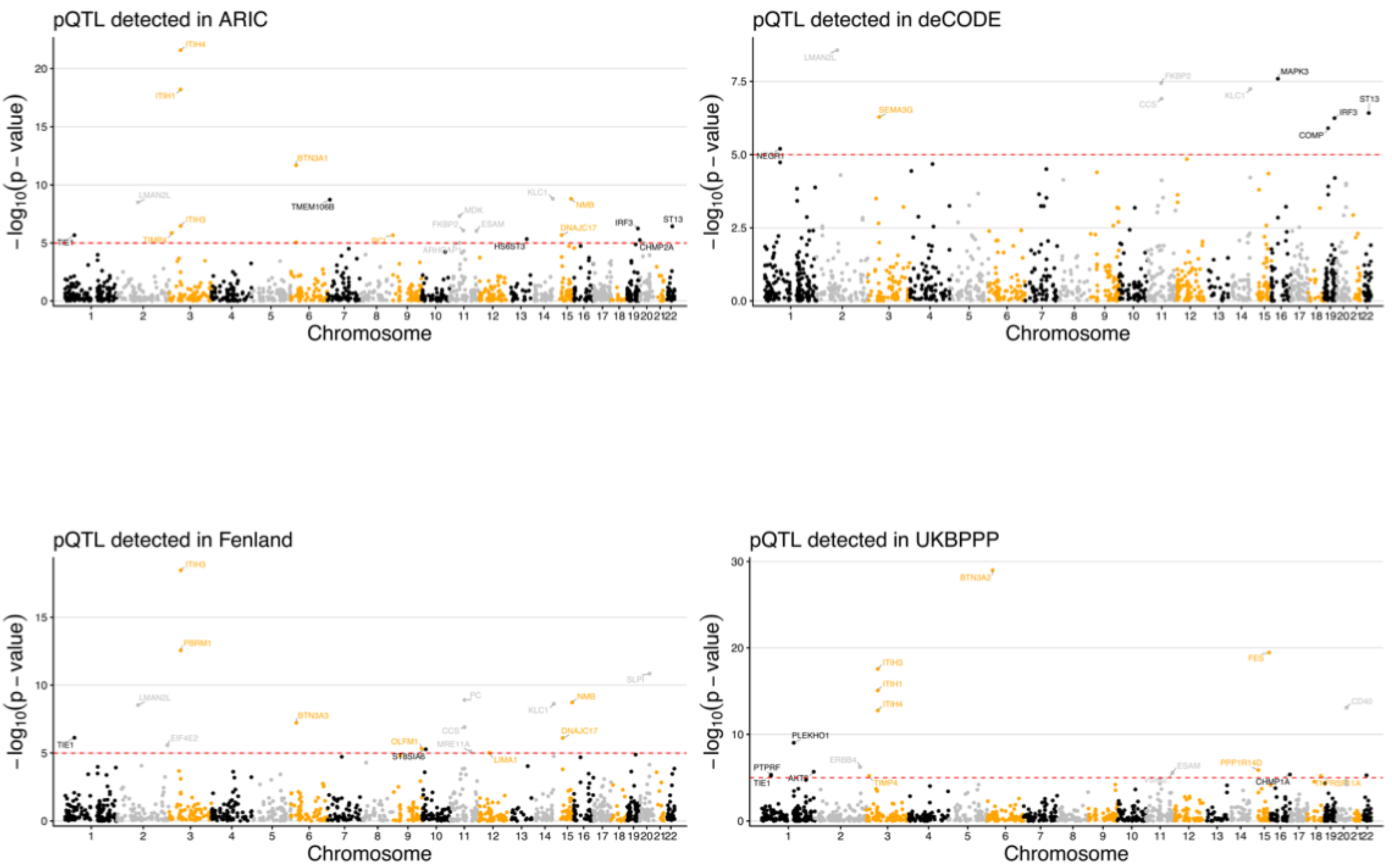


Supplementary Figure 1. Proteome-wide MR results. Manhattan plots show MR-estimated associations between circulating proteins and the common factor in the primary analyses using the IVW or Wald ratio estimator based on pQTLs identified in four studies. Each dot represents a protein, with genomic location determined by its protein-coding gene. Red dashed lines indicate the significance threshold ($P < 1x10^{-5}$).

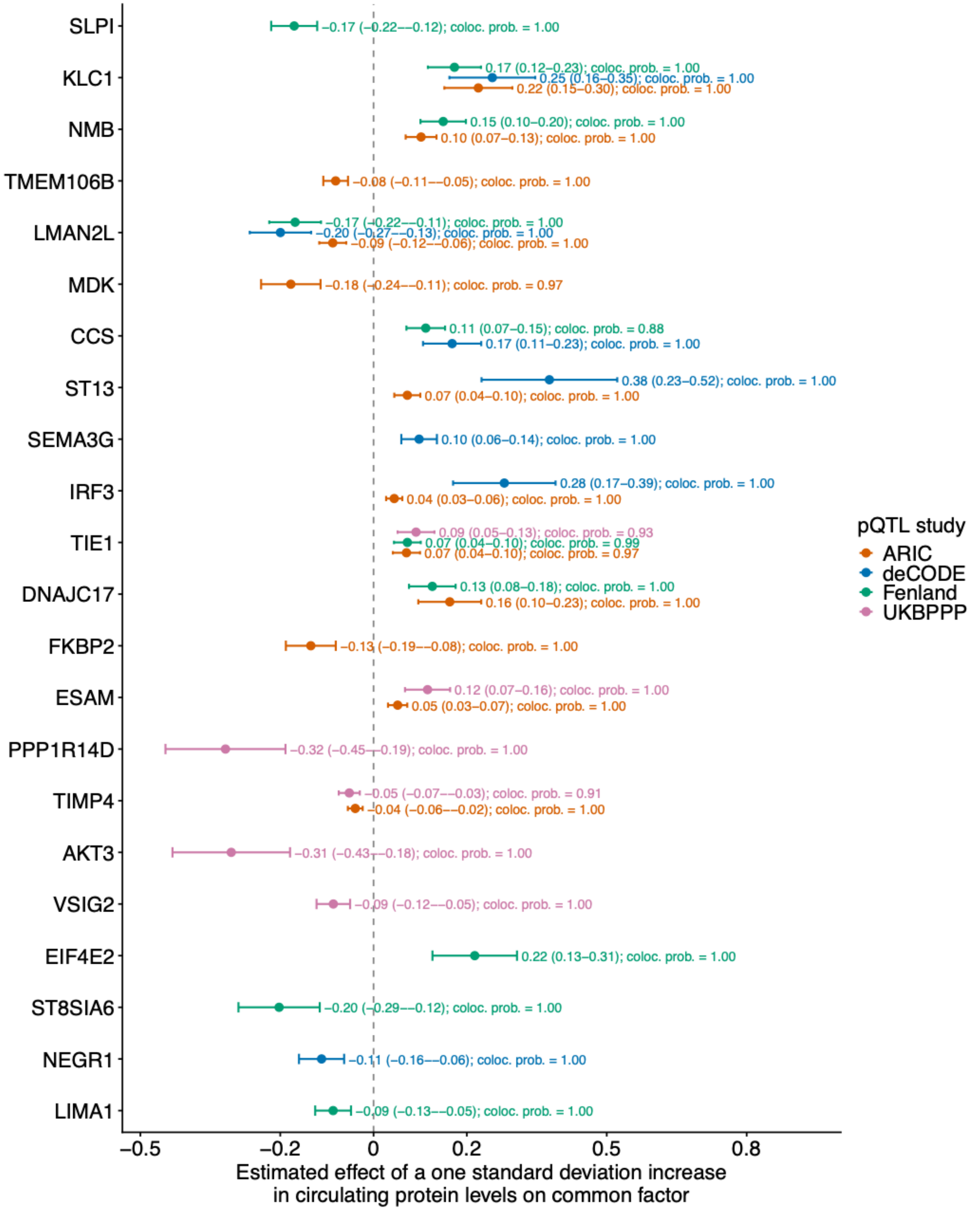


Supplementary Figure 2. Illustration of estimated associations between circulating protein levels and common factor without enrichment in brain, immune, or liver tissues. Associations that were significant in MR analyses and withstood sensitivity analyses and colocalization analyses are illustrated. Each dot denotes to the estimated effect of a protein based on a specific pQTL study, with error bars indicating 95% confidence intervals. Colocalization probabilities (coloc. prob.) are indicated. The vertical dashed line denotes the null.

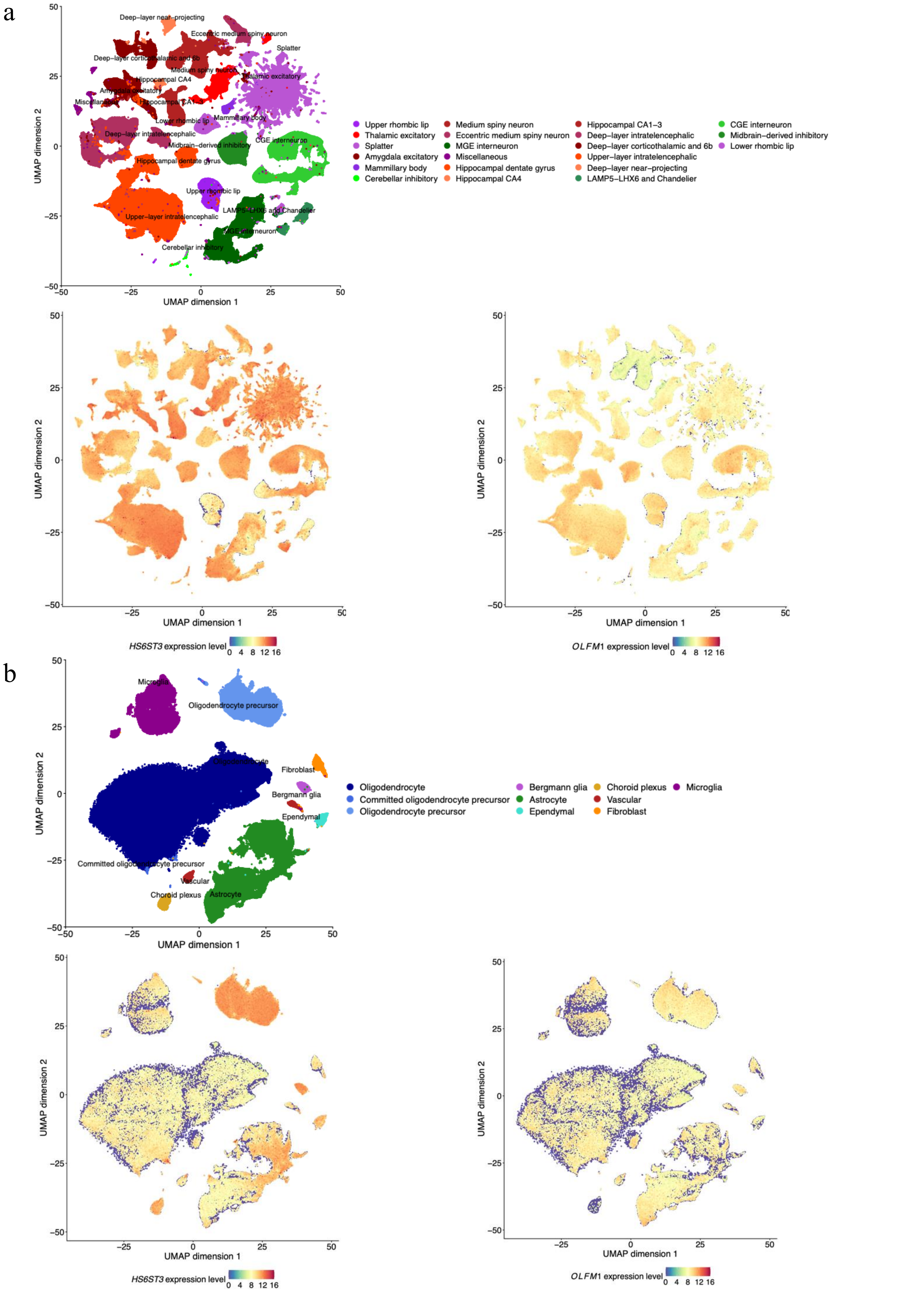


Supplementary Figure 3. Single-cell gene expression patterns of *HS6ST3* and *OLFM1* in human brain tissues. Expression patterns were visualized using uniform manifold approximation and projection (UMAP) based on transcriptomic data from more than 3 million cells. (a) Cell-type annotations and normalized gene expression levels in neuronal cell populations. (b) Cell-type annotations and normalized gene expression levels in non-neuronal cell populations.

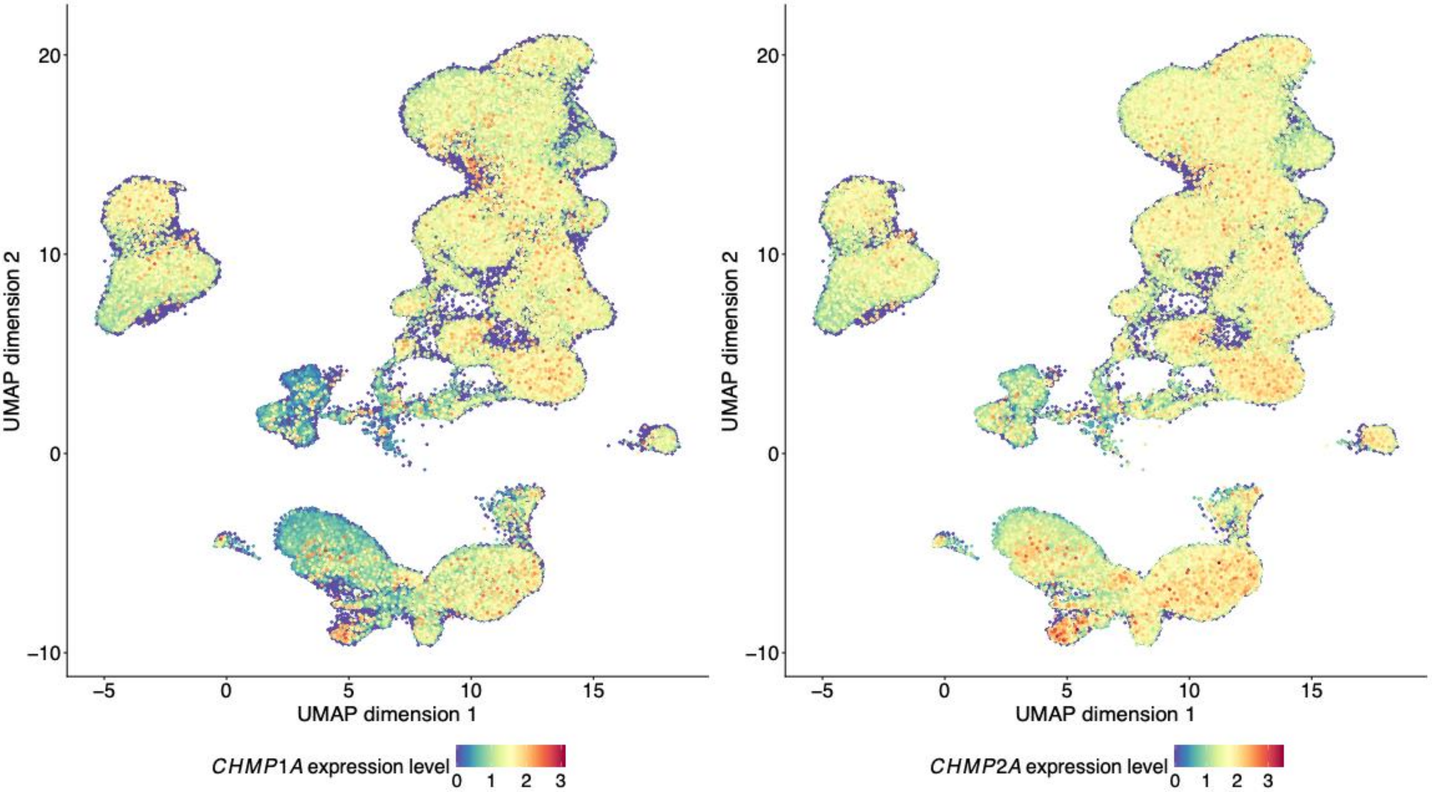


Supplementary Figure 4. Single-cell gene expression patterns of *CHMP1A* and *CHMP2A* across immune cell populations. Normalized gene expression levels are shown. Cell type annotations are illustrated in Figure 5a.

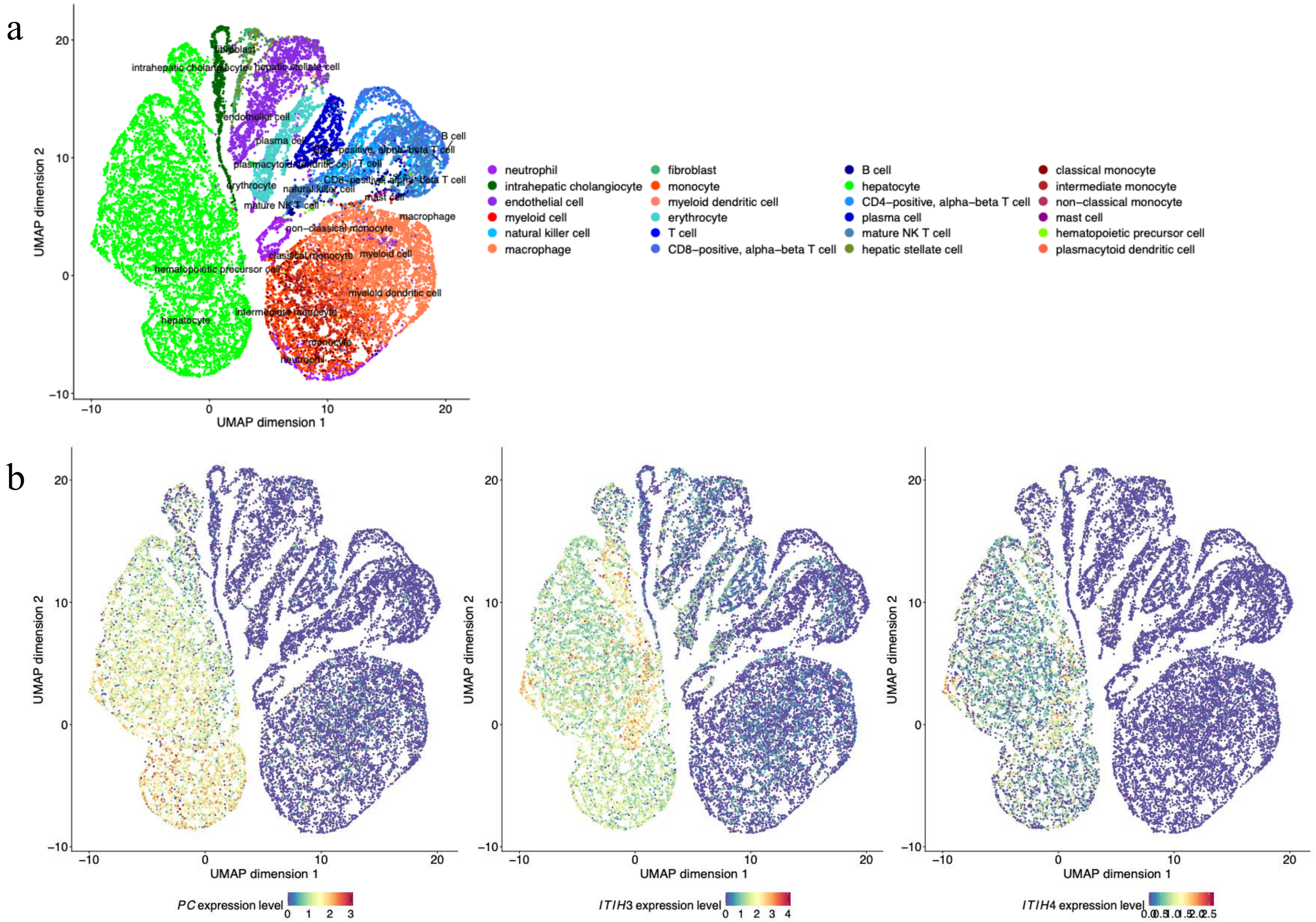


Supplementary Figure 5. Single-cell gene expression patterns of *PC*, *ITIH3* and *ITIH4* in liver. Expression patterns were visualized using uniform manifold approximation and projection (UMAP) based on transcriptomic data from approximately 10,000 cells. (a) Cell type annotations are illustrated. Each dot represents a single cell colored according to manually curated cell types. (b) Normalized gene expression levels are shown.
